# Effectiveness of Combined versus Single Circadian Interventions in Pediatric Intensive Care Units: a Systematic Review Protocol Protocol for a Systematic Review of Circadian Interventions in PICUs

**DOI:** 10.1101/2025.08.19.25334031

**Authors:** C.C. Corrotea-Maltez, C.V. Fernández-Flores, C.L. Gutierrez-Neira, A.J. Nirrian-Pérez, J. Mansilla-Muñoz, L. Bustos-González

**Affiliations:** School of Nursing, Faculty of Medicine, Universidad de Valparaíso, Angamos 655, 2540064, Viña del Mar, Chile

**Author notes:** These authors contributed equally to this work (C.C. Corrotea-Maltez, C.V. Fernández-Flores, C.L. Gutierrez-Neira, A.J. Nirrian-Pérez, J. Mansilla-Muñoz).

## Abstract

**Background:** Circadian rhythms regulate fundamental biological processes such as sleep– wake cycles, hormonal secretion, immune function, and cellular repair. In pediatric intensive care units (PICUs), these rhythms are highly vulnerable to disruption from constant artificial lighting, frequent procedures, environmental noise, and irregular feeding schedules. Critically ill children are particularly at risk due to their still-developing circadian systems, and disruption has been linked to poorer sleep quality, and delayed recovery. Although interventions such as light therapy, noise reduction, and structured sleep protocols have been explored, the comparative benefits of combining multiple circadian interventions versus applying them in isolation remain unclear.

**Objective:** This protocol outlines the methodology for a systematic review to evaluate the effectiveness of combined versus single-component circadian interventions in improving clinical outcomes, circadian alignment, sleep quality, and overall well-being in patients admitted to PICUs.

**Methods:** Following the Preferred Reporting Items for Systematic Reviews and Meta-Analyses Protocols (PRISMA-P) guidelines, we will search PubMed, Web of Science, Scopus, Embase, CINAHL, and the Cochrane Library from January 2020 to September 2025. Eligible studies will include randomized controlled trials, quasi-experimental studies, and observational designs evaluating circadian interventions among patients aged 0–18 years in PICUs. Two independent reviewers will screen records, extract data, and assess risk of bias using RoB 2 for RCTs and ROBINS-I for non-randomized studies. Narrative synthesis will be the primary analysis method; if data permit, meta-analysis will be conducted using random-effects models. Primary outcomes will include circadian measures, sleep parameters, and clinical outcomes such as length of stay and delirium incidence. Subgroup, sensitivity, and, if feasible, meta-regression analyses will be performed.

**Discussion:** Findings will inform clinical strategies to protect circadian function in critically ill children and may guide PICU environmental and care protocols.

**Registration:** This protocol is registered with the Open Science Framework (DOI: 10.17605/OSF.IO/8SPZM) and PROSPERO (CRD4201091137).

## Introduction

Chronobiology, or circadian biology, is the field that studies cyclic physiological phenomena in living organisms (1). Recent advances have expanded our understanding of the circadian clock, giving rise to Circadian Medicine, a subfield that applies chronobiological knowledge to clinical care, aiming to improve health outcomes based on circadian principles (2–5). Increasing evidence supports the view that a well-synchronized biological clock is essential for optimal health (6), and disruptions to these rhythms have been linked to a range of conditions, including mental health disorders, cardiovascular disease, and metabolic dysfunction (2).

Maintaining synchronization between the internal biological clock and environmental cues is crucial for physical and psychological well-being (7). Utilizing these strategies, the circadian clock can regulate physiological processes such as the secretion of hormones like glucocorticoids, body temperature, blood pressure, and insulin release, as well as glucose homeostasis. Additionally, food can also act as an additional *zeitgeber*, potentially entraining the peripheral clocks of the liver and pancreas (7). Proper alignment between *zeitgebers* and the central clock is fundamental to the robust and effective functioning of the internal rhythms, which in humans has been shown to promote health (6,8).

However, in modern society, individuals are frequently exposed to inappropriate or weak time cues, leading to circadian misalignment. This misalignment, caused by factors such as shift work, jet lag, or excessive artificial light exposure, has been associated with a range of adverse health outcomes including sleep disorders, depression, and cancer (9–12).

Hospitals, particularly intensive care units (ICUs), present environments that exacerbate circadian misalignment. Their 24/7 operations prioritize administrative or clinical efficiency over patient circadian rhythms (13). ICU patients often face continuous artificial light, noise, irregular procedures, and poorly timed meals, conditions that disrupt normal circadian signaling (14,15). Frequent nurse interventions and disrupted sleep-wake schedules further impair circadian alignment and delay recovery (16–18).

Pediatric intensive care units (PICUs) pose additional challenges. Infants and young children, whose circadian systems are still developing, depend heavily on environmental time cues (19,20). Circadian disruption in this population may have amplified effects, including impaired sleep, delayed recovery, and increased stress responses(21). Additionally, studies show that light and noise levels in neonatal and pediatric ICUs often exceed WHO recommendations, which may contribute to sleep fragmentation and long-term neurodevelopmental issues (22–27).

Although a growing body of literature emphasizes the importance of circadian interventions in improving patient outcomes in critical care, there remains a lack of randomized controlled trials specifically focused on the benefits of these strategies (28). This gap is particularly evident in pediatric populations in critical care. A quick search of the PubMed database using the search terms “*Circadian Rhythms AND Intervention*” retrieved a total of 23,573 studies over the past five years. In contrast, when the term *“Pediatric*” was added, the number dropped to 1,095, highlighting a notable gap in research focused on children in critical care. While some reviews have addressed circadian care in adult ICUs, no systematic review to date has synthesized the available evidence on circadian-focused nursing care strategies in PICUs. Individual circadian strategies such as light therapy and noise reduction have been explored, but there is limited evidence regarding the effectiveness of multicomponent or bundled approaches that combine these strategies. Emerging findings in adult ICU settings suggest that such integrative approaches may enhance sleep quality, reduce the incidence of delirium, and shorten hospital stays more effectively than isolated measures (29–31). However, this evidence has not been sufficiently extended to pediatric populations, where the neurodevelopmental vulnerability of children and their dependency on structured routines may influence intervention outcomes. Thus, a systematic review focusing specifically on circadian-based nursing interventions in PICUs is warranted to inform clinical practice and support the design of tailored strategies for this vulnerable population (32).

This protocol describes the methodology for a systematic review that aims to evaluate the effectiveness of multicomponent circadian interventions compared to isolated circadian strategies in enhancing clinical recovery and overall well-being among pediatric patients admitted to neonatal and pediatric intensive care units. Findings from this review are expected to inform clinical practice guidelines, guide the development of evidence-based circadian care protocols, and highlight priority areas for future research aimed at optimizing circadian alignment in critically ill children.

### Protocol Registration

The protocol for this systematic review was registered prior to study initiation with the Open Science Framework (OSF; https://doi.org/10.17605/OSF.IO/8SPZM) and the International Prospective Register of Systematic Reviews (PROSPERO; Registration No. CRD4201091137.

## Methods

### Eligibility Criteria

The eligibility criteria for this systematic review were defined in accordance with the PRISMA-P guidelines (Preferred Reporting Items for Systematic Review and Meta-Analysis Protocols) (33). The PRISMA-P checklist is available in S1 File. Criteria were structured using the PICOS framework: Population, Intervention, Comparator, Outcomes, and Study Design, to ensure clarity and methodological transparency. The inclusion and exclusion criteria are outlined below:

#### Inclusion Criteria

- Population (P): Studies involving pediatric patients (from birth to 18 years) admitted to PICUs or critical/intermediate care units, regardless of illness severity.
- Interventions (I): Circadian-based interventions aimed at enhancing patient recovery or well-being. These may include:

- Light exposure regulation
- Sleep–wake cycle optimization
- Timing of feeding
- Other strategies promoting circadian alignment.
- Comparators (C): Studies comparing circadian interventions to standard care, no intervention, or alternative circadian approaches (e.g., single vs. multicomponent interventions).
- Outcomes (O): Studies must report at least one clinical outcome related to patient recovery or well-being, including (but not limited to):

- Hospital length of stay
- Feeding tolerance
- Blood glucose regulation
- Sleep quality
- Circadian-regulated hormonal levels (e.g., cortisol, melatonin)
- Circadian regulation of physiological parameters (e.g., blood pressure, body temperature, heart rate)
- Study Design (S): Primary quantitative studies, including:

- Randomized controlled trials (RCTs)
- Quasi-experimental studies
- Observational designs (cross-sectional or longitudinal) Studies must include evaluation of clinical impact.
- Language: Publications in English, Spanish, or Portuguese.
- Timeframe: Articles published from 2020 to 2025, based on recommendations to prioritize recent advances in rapidly evolving clinical fields (34).
- Availability: Full-text articles published in peer-reviewed scientific journals.

#### Exclusion Criteria

- Study type: Systematic, scoping, or narrative reviews, and qualitative studies.
- Population focus: Studies focusing exclusively on healthcare providers (e.g., nurses or physicians), without reporting patient data.
- Language: Publications in languages other than English, Spanish, or Portuguese (e.g., French, Italian).
- Access and format: Articles not available in full text.
- Scope: Studies that focus solely on environmental factors (e.g., noise, lighting, routines) without reporting direct clinical outcomes in pediatric patients.
- Outcome focus: Studies reporting only staff outcomes or that do not assess patient-specific clinical effects.

### Information Sources

A comprehensive literature search will be conducted using the following electronic databases: PubMed, Scopus, Web of Science, CINAHL, LILACS, SciELO, Elsevier, and EBSCOhost. These databases were selected to provide broad coverage of published studies, encompassing both global and Latin American research contexts.

The search will be limited to studies published between January 2020 and September 2025, with no geographical restrictions. Eligible publications must be in English, Spanish, or Portuguese. Grey literature and preprints will be excluded to ensure the inclusion of peer-reviewed sources only.

In addition to the electronic search, the reference lists of all included studies and relevant systematic reviews will be manually screened to identify additional eligible studies not captured in the initial database search.

### Search Strategy

The search strategy will combine controlled vocabulary from MeSH (Medical Subject Headings) and DeCS (Health Sciences Descriptors), with relevant free-text terms. The core search concepts will include:

- English Terms: Circadian rhythm, Intensive Care Units, Pediatric, Nursing care
- Spanish Terms: Ritmo circadiano, Unidades de Cuidados Intensivos Pediátricos, Atención de enfermería.

Boolean operators (e.g., “Circadian Rhythm” AND “Intensive Care Units, Pediatric” AND “Nursing Care”) will be applied to refine results. Each database will have a search strategy adapted to its indexing system, syntax, and available filters. The full, database-specific search strings, including any applied limits, are provided in S2 File.

### Study Records

#### Data Management

All records retrieved from the database searches will be imported into EndNote Web for initial duplicate detection and removal. The deduplicated dataset will then be exported to Covidence, a systematic review management platform recommended in the Cochrane Handbook for Systematic Reviews of Interventions (34) Covidence will be used for additional duplicate removal, as well as for screening, study selection, and record management. This platform will serve as the central repository for organizing, tracking, and documenting all decisions made during the review, thereby ensuring methodological transparency and reproducibility.

#### Selection Process

The study selection will be conducted in three stages: (1) title screening, (2) abstract screening, and (3) full-text review. Each stage will be performed independently by four reviewers from the research team. Discrepancies will be discussed among reviewers, and consensus will be sought through deliberation. If consensus cannot be reached, a fifth reviewer will adjudicate the decision. All screening will be managed in Covidence, which supports blinded assessments and conflict resolution workflows. Eligibility will be determined according to the predefined inclusion and exclusion criteria established for this review.

#### Data Collection Process

Data from the included studies will be extracted using a standardized form developed specifically for this review. Prior to full implementation, the form will be piloted on a subset of eligible studies to ensure clarity, completeness, and inter-reviewer consistency. Data extraction will be performed independently by two reviewers to minimize bias. Any discrepancies will be resolved through discussion; if consensus is not reached, a third reviewer will adjudicate.

The following data items will be collected:

- General study characteristics: title, authors, year of publication, country or region, language, and source of funding (if available).
- Study design and methodology: type of study (e.g., randomized controlled trial, quasi-experimental), sample size, duration of follow-up, and methodological details relevant to the risk of bias assessment.
- Population characteristics: age range, mean age (if reported), sex distribution, clinical condition or diagnosis, and inclusion/exclusion criteria.
- Intervention characteristics: type of circadian intervention (e.g., dynamic lighting, environmental noise reduction, scheduled feeding, sleep promotion bundles), mode of delivery, timing, duration, and frequency.
- Comparator(s): description of control group, standard care, or alternative interventions.
- Outcomes: main outcomes related to circadian function and clinical recovery, such as length of hospital stay, sleep quality, feeding tolerance, blood glucose regulation, body temperature, hormonal levels (e.g., melatonin, cortisol), delirium incidence, and physiological indicators of circadian rhythm (e.g., heart rate variability, blood pressure).
- Results and effect estimates: effect sizes, measures of variability (e.g., confidence intervals), and statistical significance (p-values).
- Feasibility and implementation details: adherence to the intervention, staff involvement, and reported barriers or facilitators (when available).

All data will be managed in Covidence, which supports collaborative extraction, transparent tracking of decisions, and export for subsequent analysis and synthesis.

### Data Items

The following variables will be extracted from each included study:

- General study characteristics: Title, authors, publication year, country/region, language, and funding source (if available).
- Study design and methodology: Study type, sample size, follow-up duration, and methodological notes relevant to risk of bias assessment.
- Population characteristics: Age, sex, clinical condition/diagnosis, and inclusion/exclusion criteria.
- Intervention details: Type of circadian intervention (e.g., dynamic lighting, noise reduction, scheduled feeding, multicomponent bundles), delivery mode, timing, frequency, and duration.
- Comparator(s): Description of control or standard care, or alternative circadian strategy.
- Outcomes: Clinical and circadian outcomes (e.g., sleep quality, hormonal levels such as melatonin/cortisol, feeding tolerance, length of hospital stay, delirium incidence, physiological circadian markers).
- Results and effect estimates: Effect sizes, confidence intervals, and p-values.
- Feasibility and implementation: Adherence, staff roles, and reported barriers or facilitators (if available).

If multiple timepoints are reported, all relevant timepoints will be extracted. When data are missing or unclear, study authors will be contacted for clarification, and any assumptions made will be documented transparently.

### Outcomes and Prioritization

#### Primary outcome

The primary outcome of this systematic review is the effectiveness of circadian interventions on clinical recovery in pediatric intensive care patients. This will be operationalized primarily through:

- Length of hospital stay, which is commonly used as an indicator of PICU recovery and resource utilization (35)
- Sleep quality, measured via validated instruments or physiological indicators, as circadian-related outcomes frequently disrupted in PICU patients (21,36)

#### Secondary outcomes

Secondary outcomes include:

- Physiological circadian markers (e.g., melatonin, cortisol, body temperature, heart rate variability, blood pressure);
- Delirium incidence during hospitalization, which has been linked to circadian rhythm dysregulation in critically ill children (21,37)
- Feeding tolerance and metabolic regulation (e.g., glycemic control);
- Feasibility and adherence, including reported barriers and facilitators;
- Patient comfort and emotional well-being, when available.

Outcomes were prioritized based on clinical significance, frequency of reporting, and relevance to circadian function in critically ill pediatric populations. Data will be captured across all reported timepoints to assess both short-term and longer-term effects.

### Risk of Bias and Study Quality Assessment in Individual Studies

To ensure a robust evaluation of internal validity, methodological rigor, and reporting quality, we will conduct a multi-layered assessment tailored to the design of each included study.

#### Randomized Controlled Trials (RCTs)

Risk of bias will be assessed using the Risk of Bias tool, version 2 (RoB 2), developed by the Cochrane Collaboration (38). This tool evaluates domains such as:

1. Randomization process
2. Deviations from intended interventions
3. Missing outcome data
4. Measurement of outcomes
5. Selection of the reported result

Methodological quality will be appraised using the CASPe checklist for RCTs (39).

#### Non-Randomized Studies

Risk of bias will be evaluated using the Risk Of Bias In Non-randomized Studies of Interventions (ROBINS-I) tool (40), which examines potential biases from:

1. Confounding
2. Participant selection
3. Classification of interventions
4. Deviations from intended interventions
5. Missing data
6. Outcome measurement
7. Selection of the reported result.

Methodological quality will be assessed with the CASPe checklists for cohort and case– control studies (39). Reporting quality will be evaluated using the STROBE checklist (41), enabling identification of incomplete or inconsistent reporting that could influence interpretation.

#### Assessment Process

Two independent reviewers will conduct all assessments. Disagreements will be resolved through discussion, and when necessary, a third reviewer will adjudicate. Assessment results will be presented in both tabular and narrative formats. Although studies will not be excluded solely based on high risk of bias or poor reporting quality, these evaluations will be used to:

- Inform sensitivity analyses.
- Contextualize the strength, consistency, and applicability of the evidence.

### Data Synthesis

Given the anticipated heterogeneity in study designs, participant populations, interventions, and outcome measures, the primary approach will be a narrative synthesis. Findings from included studies will be organized thematically, with results grouped by:

- Type of circadian intervention (multicomponent vs. single-component).
- Clinical outcomes (e.g., length of hospital stay, sleep quality, hormonal markers, delirium)
- Implementation characteristics (e.g., adherence, feasibility, contextual barriers/facilitators)

Special emphasis will be placed on comparing the effectiveness of multicomponent circadian interventions versus isolated strategies in improving clinical recovery and overall well-being among pediatric patients in PICUs.

Studies will be further grouped according to:

- Intervention characteristics (e.g., light modulation, noise reduction, feeding schedules)
- Participant characteristics (e.g., age range, diagnosis, illness severity)
- Outcome domains (physiological, behavioral, and patient-centered outcomes)

Patterns, inconsistencies, and contextual factors will be examined to explore the potential added value of multicomponent interventions over single-component approaches. Where possible, we will consider whether differences in results can be explained by study setting, population, or intervention design.

If subsets of studies are found to be sufficiently homogeneous in terms of design, interventions, and outcome measures, a meta-analysis will be conducted using RevMan software (42), in line with Cochrane Handbook recommendations (34). Effect measures will be selected according to the type of outcome: mean difference (MD) or standardized mean difference (SMD) for continuous data, and risk ratio (RR) for dichotomous outcomes. Fixed- or random-effects models will be applied based on the degree of heterogeneity, assessed through the *I²* statistic and, where relevant, Kendall’s τ.

If a meta-analysis is feasible, additional analyses will be undertaken, including sensitivity analyses to evaluate the influence of study quality, risk of bias, and inclusion criteria. Subgroup analyses will be planned to explore differences based on intervention type, patient age group, or illness severity. Where possible, meta-regression will be considered to assess the impact of continuous moderators (e.g., intervention duration, light intensity).

Where meta-analysis is not possible, findings will be integrated narratively, ensuring transparent reporting of data handling, synthesis methods, and limitations.

### Meta-bias(es)

To address potential meta-biases, we will evaluate the risk of selective outcome reporting and publication bias across included studies. When study protocols or trial registrations (e.g., ClinicalTrials.gov, WHO ICTRP) are available, we will compare prespecified outcomes with those reported in the published results to detect discrepancies. In cases where protocols are not accessible, we will assess consistency between the methods and results sections to identify potential selective reporting (34).

If sufficient studies (≥10) are available for a meta-analysis, we will generate funnel plots to visually examine small-study effects or publication bias in accordance with the Cochrane Handbook for Systematic Reviews of Interventions (34). Egger’s regression test will be used to statistically evaluate funnel plot asymmetry (43), and where asymmetry is detected, the trim-and-fill method will be applied as a sensitivity analysis to estimate the potential impact of missing studies (44).

When fewer than 10 studies are available, or substantial heterogeneity precludes meaningful statistical assessment, formal bias testing may not be feasible. In such cases, we will consider qualitative signals of bias, such as overrepresentation of statistically significant findings, absence of null-effect studies, or outcome switching. All findings related to potential reporting or publication biases will be documented transparently and considered in the interpretation of the overall results.

### Confidence in Cumulative Evidence

We will assess the overall confidence in the cumulative evidence for each primary and secondary outcome using the Grading of Recommendations Assessment, Development, and Evaluation (GRADE) framework (45). Given that this review will include both randomized controlled trials (RCTs) and non-randomized studies (NRS) comparing combined versus isolated circadian interventions in pediatric intensive care units (PICUs), the approach will be adapted to account for the methodological diversity.

In PICU research, conducting RCTs on circadian-based interventions often involves ethical, logistical, and methodological challenges. For example, assigning critically ill children to suboptimal light or noise conditions may not be ethically permissible, and many interventions are implemented at the unit level, limiting the feasibility of individual randomization. High-quality NRS can therefore provide valuable insights, particularly regarding real-world implementation of environmental or behavioral interventions. Including NRS in this review will enhance the ecological validity of findings while capturing a broader range of evidence. All NRS will be appraised using the ROBINS-I tool(40) and their contribution to the cumulative evidence will be assessed using the GRADE approach adapted for observational designs (46)

For RCTs, the standard GRADE methodology will be applied, assessing:

1. Risk of bias, using the RoB 2 tool (38)
2. Inconsistency, variability in results across studies
3. **I**ndirectness, relevance of population, intervention, comparator, and outcome
4. Imprecision, confidence intervals and sample sizes
5. Publication bias, suspected through patterns in reported results.

For NRS, certainty of evidence will start at “low” and be downgraded or upgraded based on:

1. Risk of bias (ROBINS-I)
2. Inconsistency, indirectness, imprecision, and publication bias
3. Magnitude of effect
4. Dose–response gradient
5. Likelihood that residual confounding would change the observed effect.

When both RCTs and NRS report on the same outcome, we will assess coherence between designs. If findings are consistent, confidence will be strengthened; if results differ, RCT data will be prioritized unless NRS offer more relevant or robust contextual information.

All evidence profiles will be summarized in Summary of Findings (SoF) tables created with GRADEpro GDT (47), reporting effect sizes, participant numbers, number of studies, and certainty ratings (high, moderate, low, very low). For NRS, the STROBE checklist (41) will support transparent reporting and interpretation.

### Study Status and Timeline

As this project is a systematic review, no participant recruitment will be conducted. Data collection (i.e., literature search and study selection) is expected to be completed by September 9, 2025. Based on the available evidence in adult ICU populations, the anticipated results are that multicomponent circadian interventions are more effective than single-component strategies, although adherence tends to be low.

### Ethics and Dissemination

This review will synthesize data from previously published studies and will not involve the collection of primary data from human participants. Therefore, ethical approval is not required. The findings will be disseminated through peer-reviewed publication and presentations at relevant conferences.

### Updates to study protocol

Any amendments to this review protocol will be documented in detail, appended as supplementary material to the final manuscript, and reflected in the updated records of both the Open Science Framework (OSF; DOI: 10.17605/OSF.IO/8SPZM) and PROSPERO (Registration ID: CRD4201091137).

## Discussion

This protocol outlines a systematic review designed to evaluate the effectiveness of circadian-based interventions in improving clinical recovery among pediatric patients admitted to intensive care units. While circadian rhythm disruption is a recognized concern in critically ill populations, particularly in children whose developmental physiology makes them more susceptible to environmental disturbances, evidence synthesis in this area remains scarce and fragmented. This review will address an important gap by examining both clinical outcomes (e.g., hospital length of stay, sleep quality) and physiological indicators (e.g., hormonal levels, heart rate variability) in response to interventions aimed at circadian alignment.

The inclusion of both RCTs and high-quality NRS will ensure a more comprehensive appraisal of the available evidence. This is especially relevant given the ethical and logistical barriers that often limit RCT feasibility in PICU settings. By adopting the GRADE framework for both study types, this review will provide a structured evaluation of the certainty of evidence, enabling clear guidance for clinical and research applications.

Findings will also have important implications for clinical practice. Circadian-focused strategies, such as dynamic lighting systems, noise reduction protocols, and time-based feeding interventions, may offer low-cost, scalable approaches to improving patient outcomes. However, their implementation in high-intensity clinical environments requires careful consideration of feasibility, staff engagement, and unit-specific constraints. Understanding facilitators and barriers will be critical for translating evidence into practice.

### Strengths and Limitations

A key strength of this protocol is its adherence to internationally recognized methodological guidelines (PRISMA-P), including transparent eligibility criteria and a pre-registered protocol in both OSF and PROSPERO. The comprehensive search strategy incorporates both MeSH and DeCS terms, increasing the likelihood of capturing relevant studies across global and Latin American contexts. Furthermore, the use of validated tools (RoB 2, ROBINS-I, CASPe, STROBE) for risk of bias and quality assessment will enhance the reliability of findings.

However, some limitations should be acknowledged. Restricting inclusion to studies published in English, Spanish, and Portuguese may lead to language bias, potentially excluding relevant evidence in other languages. The anticipated heterogeneity of interventions, patient populations, and outcome measures may limit the feasibility of conducting a meta-analysis, with narrative synthesis likely to be the primary analytical approach. Finally, as with all systematic reviews, the quality of conclusions will depend on the methodological rigor and reporting quality of the included studies.

## Data Availability

No datasets were generated or analysed during the current study. All relevant data from this study will be made available upon study completion.

## Acknowledgments

We thank the members of the School of Nursing at the University of Valparaíso, particularly Dr. (c) Lorena Bettancourt and Dr. (c) Cibeles González, coordinators of the Project I and II courses, for their methodological guidance and feedback.

## Author Contributions

- Conceptualization: L. Bustos-González
- Methodology: L. Bustos-González, C.C. Corrotea-Maltez, C.V. Fernández-Flores, C.L. Gutiérrez-Neira, A.J. Nirrian-Pérez, J. Mansilla-Muñoz
- Formal analysis: C.C. Corrotea-Maltez, C.V. Fernández-Flores, C.L. Gutiérrez-Neira, A.J. Nirrian-Pérez, J. Mansilla-Muñoz
- Data curation: C.C. Corrotea-Maltez, C.V. Fernández-Flores, C.L. Gutiérrez-Neira, A.J. Nirrian-Pérez, J. Mansilla-Muñoz
- Investigation: C.C. Corrotea-Maltez, C.V. Fernández-Flores, C.L. Gutiérrez-Neira, A.J. Nirrian-Pérez, J. Mansilla-Muñoz
- Writing-original draft: L. Bustos-González
- Writing-review & editing: All authors
- Supervision: L. Bustos-González

## List of Abbreviations

CASPe: Critical Appraisal Skills Programme Español
GRADE: Grading of Recommendations Assessment, Development and Evaluation
NRS: Non-Randomized Studies
OSF: Open Science Framework
PICU: Pediatric Intensive Care Unit
PRISMA-P: Preferred Reporting Items for Systematic Review and Meta-Analysis Protocols
RCT: Randomized Controlled Trial
ROBINS-I: Risk Of Bias In Non-randomized Studies - of Interventions
RoB 2: Cochrane Risk of Bias tool version 2
SoF: Summary of Findings
STROBE: Strengthening the Reporting of Observational Studies in Epidemiology

## Supporting information

**S1 File. PRISMA-P (Preferred Reporting Items for Systematic review and Meta-Analysis Protocolos) 2015 checklist: Recommended items to address in a systematic review protocol.**

**S2 File. Search strategy.**

